# Psychological factors associated with instrumental activities of daily living disability in older adults with moderate to severe knee osteoarthritis

**DOI:** 10.1101/2021.09.10.21262871

**Authors:** Keigo Nanjo, Takashi Ikeda, Naoko Nagashio, Tomoko Sakai, Tetsuya Jinno

## Abstract

**BACKGROUND:** The population of older adults with knee osteoarthritis (OA)-related disabilities is increasing globally. However, studies regarding instrumental activities of daily living (IADL) in older adults with knee OA are limited.

**OBJECTIVE:** This study investigated the psychological factors associated with IADL disability in older adults with moderate to severe knee OA.

**METHODS:** A cross-sectional study was conducted on 179 patients with knee OA aged ≥ 65 years. The six-item short form of the Pain Catastrophizing Scale (PCS-6), the four-item short form of the Pain Self-Efficacy Questionnaire (PSEQ-4), and the fifteen-item Geriatric Depression Scale (GDS-15) were used to assess psychological factors. The participants were divided into IADL-disabled and non-disabled groups. Binary logistic regression analyses were performed with the IADL disability status as the dependent variable. The PCS-6, PSEQ-4, and GDS-15 tools were included as independent variables in the logistic regression model.

**RESULTS:** Of the total participants, 88 (49.1%) showed disability in conducting IADL. PSEQ-4 (odds ratio =0.90, 95%; confidence interval=0.82–0.99, p=0.02) was a significant independent variable among all psychological factors.

**CONCLUSION:** Our study showed the importance of assessing self-efficacy using the PSEQ-4 in relation to IADL disability in older adults with moderate to severe knee OA.

## 1. Introduction

Knee osteoarthritis (OA) is a prevalent age-related chronic condition and a common cause of disabilities limiting daily activities in older adults [1]. Since the population of older adults with knee OA-related disabilities is increasing globally [2], healthcare professionals need effective countermeasures to address this concern.

Instrumental activities of daily living (IADL) are essential elements in the independent lives of older adults. It is defined as independent functioning in a given environment and includes activities such as meal preparation, shopping, commutation, financial management, and performance of other household chores [3]. Meanwhile, basic activities of daily living (BADL) include basic self-care activities such as bathing, dressing, eating, and other indoor activities [4]. IADL require more complex functioning than BADL. As IADL disability in older adults influences health-related quality of life [5], all-cause mortality [6], and decline of cognitive function [7], assessments and interventions for IADL disability are important for this population.

Disabilities in patients with knee OA are affected by factors such as gait speed, knee muscle strength, and pain [8, 9]. Psychological factors, such as pain catastrophizing [10–12], self-efficacy [12–14], and depressive symptoms [14–16], have also been reported among disabled patients with knee OA. A meta-analysis has shown the importance of psychological intervention for disability in patients with knee OA [17].

However, most studies of psychological factors and disability among patients with knee OA [10–16] have treated disability as a concept that combines BADL and IADL. It is well known that disabilities in older adults progress hierarchically [18]; IADL disability occurs before BADL disability [19]. Especially among older adults with knee OA, countermeasures should be taken at an early stage to prevent an incident or deterioration of disabilities. In particular, older adults with moderate to severe knee osteoarthritis are more likely to develop disabilities in their daily lives than healthy older adults [20]. Therefore, investigating specific factors related to IADL disability in older adults with moderate to severe knee OA is useful information to develop effective interventions. To our knowledge, there are limited studies of IADL disability among older adults with knee OA [16, 21]; only our previous study has specified that gait speed and pain intensity are related to IADL disability in older adults with knee OA [22]. Psychological interventions are recommended for such disabilities [17]; however, which specific psychological factors are associated with IADL disability remain unclear.

This study aimed to determine the psychological factors associated with IADL disability in older adults with moderate to severe knee OA. We hypothesized that psychological factors such as pain catastrophizing, self-efficacy, and depressive symptoms are independently associated with IADL disability, even when adjusted for demographics, motor functions, and pain intensity as confounders.

## 2. Materials and Methods

### 2.1 Participants

Patients with knee OA scheduled for primary unilateral total knee arthroplasty (TKA) or unicompartmental knee arthroplasty (UKA) were eligible for this study. The inclusion criteria were those aged ≥65 years and with a diagnosis of knee OA based on the clinical guidelines: knee pain for >3 months and Kellgren-Lawrence score (KL score) of ≥2 [23]. Conversely, the exclusion criteria were those diagnosed with rheumatoid arthritis, knee OA after trauma, dementia (mini-mental state examination score <23), BADL disability (Barthel index <100), a history of lower extremity surgery, serious pathologies (e.g., cancer during treatment), and neurological findings (e.g., motor paralysis) that could influence test performance. Additionally, patients who intended to get TKA or UKA on their other knee were excluded to eliminate the effects of pain in the contralateral knee. A total of 357 patients were identified at the start of the study. Of them, 166 were excluded due to a history of TKA or UKA on the other knee (n=106), history of lower extremity surgery except knee joint (n=32), rheumatoid arthritis (n=13), Barthel index <100 (n=9), mini-mental state examination score <23 (n=3), or knee OA after trauma (n=3). A total of 191 patients were invited to participate in this study, 12 of whom refused; finally, 179 patients were included.

### 2.2 Study design

This study used a cross-sectional design, and its conducting and reporting were guided by the STROBE guidelines [24]. Sample recruitment was conducted at Shonan Kamakura General Hospital from May 17, 2019 to May 30, 2021. All participants provided informed consent before the study began. The study was conducted in accordance with the Declaration of Helsinki and was approved by the research ethics boards of the Tokushukai Group Ethics Committee (No. TGE01198-024). All participants were given a two- to three-month waiting period after deciding to undergo TKA or UKA, and physical therapists taught them self-exercises (e.g., muscle strength and range of motion [ROM] exercises). All measurements were evaluated by well-trained physical therapists one month before the surgery.

### 2.3 Outcome measures

#### 2.3.1 IADL status

IADL status was assessed based on six activity items (preparing food, shopping, housekeeping, doing laundry, using transportation, and handling finances) using the IADL scale proposed by Lawton and Brody [3]. Taking medication and using a telephone were excluded, as it was expected that the condition of knee OA would not affect these items. Participants were asked to rate their abilities to perform these IADL activities by “able,” “need help,” or “unable.” Based on previous cross-sectional reports [25, 26], we defined those participants who opted for “need help” or “unable” to perform for at least one item as “disabled”; otherwise, they were defined as “non-disabled.”

#### 2.3.2 Psychological factors

Pain catastrophizing, self-efficacy, and depressive symptoms were assessed as psychological factors based on previous reports that discussed the relationship between disability and psychological factors in patients with knee OA, as they show strong relation to disability even when adjusted for other confounders such as demographic factors and pain intensity. [10–16]

Pain catastrophizing was assessed using the Japanese version of the six-item short form of the Pain Catastrophizing Scale (PCS-6) [27]. The Pain Catastrophizing Scale consists of subscales related to magnification, rumination, and helplessness [28]. PCS-6 has the same properties as the original version, correlates to pain intensity assessed by numerical rating scales (*r*=0.30, *p*<0.001) [29], and has good internal consistency (Cronbach’s alpha=0.90) [27]. Participants were asked to rate the degree to which they experienced each of the six thoughts and feelings when experiencing pain on a scale from 0 (“not at all”) to 4 (“all the time”). The total PCS-6 score ranges from 0 to 24, with higher scores indicating higher levels of pain catastrophizing.

Self-efficacy was assessed using the Japanese version of four-item short form of the Pain Self-Efficacy Questionnaire (PSEQ-4) [30]. The Pain Self-Efficacy Questionnaire is a 10-item self-report questionnaire used to assess self-efficacy in individuals with chronic pain [31]. The PSEQ-4 has the same properties as the original version [30], correlates pain intensity assessed by numerical rating scales (*r*=-0.35, *p*<0.001), and has good internal consistency (Cronbach’s alpha=0.90) [29]. The PSEQ-4 consists of four questions, and participants were asked how confident they were to perform the given activities despite pain, on a scale of 0 (“not at all confident”) to 6 (“completely confident”). The PSEQ-4 scores range from 0 to 24, with higher scores indicating more confidence in performing the given activities despite pain.

Depressive symptoms were assessed using the Japanese version of the fifteen-item Geriatric Depression Scale (GDS-15) [32]. The GDS-15 can screen depression (area under the curve of the receiver characteristic operating curve=0.96) and has good internal consistency (Cronbach’s alpha=0.83) [32]. The participants answered with a “yes” or “no” response. The total score was calculated from 0 to 15, with higher scores indicating more depressive symptoms.

### 2.4 Confounders

We assessed gait speed and pain intensity as confounders for outcome measures, referring to previous studies that report factors related to IADL disability in older adults [22,25,26,33]. Knee muscle strength and joint ROM were also assessed as confounders, since they are considered specific factors related to disability in patients with knee OA [34]. Based on previous studies regarding IADL disability and gait speed [22,26,33], usual gait speed (UGS) was measured using a 5-meter gait test [35]. If participants use a cane on a daily basis, we permitted using it during measurement.

The Japanese version of the pain subscale of the Knee injury and Osteoarthritis Outcome Score (KOOS-pain) was used for the representative index of pain intensity [36]. The KOOS-pain consists of nine questions. Participants were asked about their condition one week before the evaluation date. Standardized answer choices were provided, and each question was assigned a score ranging from 0 to 4. A normalized total score of 0 to 100 was calculated, and higher scores indicated that patients reported less pain. The KOOS-pain correlates with the body pain subscale of the short form-36 health survey (*r*=0.67, *p*<0.01) and has good internal consistency (Cronbach’s alpha=0.90) [36].

Isometric knee extension strength (IKES) was measured using a handheld dynamometer (μ-tus F-1, Anima Corporation, Tokyo, Japan) [37]. Knee extension and flexion ROM were measured using a goniometer [38]. The knee scheduled for surgery was defined as the affected side, and the opposite side was defined as the unaffected side. IKES and knee ROM measurements were conducted on both knees. In addition to these factors, age, sex, body mass index, Charlson comorbidity index (CCI) [39], and KL score of both knees were obtained from clinical records and used as confounders.

### 2.5 Statistical analysis

The participants were divided into two groups according to their IADL status: those who answered “need help” or “unable” to perform on at least one item were assigned to the IADL disabled group, and those who answered “able” in all items were assigned to the IADL non-disabled group. All outcome measures and confounders were compared between the two groups. The chi-square test was used for categorical variables, and Student’s *t*-test and Mann–Whitney *U* test were used for normally distributed and non-normally distributed variables, respectively. In the IADL disabled group, the proportion of each item of IADL disability was shown.

Which psychological factor can be associated with IADL disability was determined using a binary logistic regression model with IADL status as the dependent variable (disabled= 1 or not= 0). As previous reports have shown that age, sex [40], gait speed [22,26,33], and pain intensity [22, 25] are factors related to IADL disability in older adults, these variables were included as confounders. In addition, other confounders that had *p-*values <0.05 in the two groups of comparison were added to the logistic regression model as confounders. As the first step, PCS-6, PSEQ-4, and GDS-15 were included as the independent variable in each model (Mode1-3) separately. Furthermore, all psychological factors (e.g., PCS-6, PSEQ-4, GDS-15) were included as the independent variable simultaneously (Model 4) because we consider them to confound with each other. All analyses were performed using R version 4.0.3 (Foundation for Statistical Computing, Vienna, Austria) for all tests; a *p*-value <0.05 was considered statistically significant.

To determine the sample size for the logistic regression analysis, the number of cases (N) =10 k/p are needed, where k is the number of independent variables as covariates and p is a defined ratio of responders to non-responders at the follow-up points [41]. To avoid overfitting and comply with the recommendations of Vittinghoff and McCulloch [42], we assumed that we would arrive at nine factors out of all the variables in the measurements and demographics. In a study involving older adults with joint pain (n=407), 60.9% of participants had IADL disability [25]. Assuming that the ratio p of disabled to non-disabled is 1:1.5, the minimum sample size in this study was calculatedLJas 10×9/0.6=150.

## 3. Results

All participants completed all assessments, and all confounders were collected from their medical records. Participants characteristic are presented in Table 1. Of all participants, 88 (49.1%) had IADL disabilities. Sixty-one (69.3%) showed disability in shopping, and 47 (53.4%) showed disability in using transportation. (Table 2).

**Table 1.** Baseline characteristics of the study population.

| Variable | ALL (n= 179) |
| --- | --- |
| Women; number (%) | 141 (78.8) |
| Age; mean±SD | 76.5 ± 5.4 |
| Body mass Index(kg/m<sup>2</sup>); median (IQR) | 23.1 (21.1–26.2) |
| Family status (living alone); number (%) | 36 (20.1) |
| CCI; median (IQR) | 0 (0–1.0) |
| KL score on affected side |  |
| □; number (%) | 19 (10.6) |
| □; number (%) | 101 (56.4) |
| □; number (%) | 59 (33.0) |
| KL score on unaffected side |  |
| □; number (%) | 62 (34.6) |
| □; number (%) | 82 (45.8) |
| □; number (%) | 35 (19.6) |

CCI, Charlson comorbidity index; IQR, interquartile range; KL score,
Kellgren-Lawrence score

**Table 2.** Content of IADL disability in IADL disabled group (n=88)

| Item | Number (%) |
| --- | --- |
| Preparing food | 33 (37.5) |
| Shopping | 61 (69.3) |
| Housekeeping | 40 (45.5) |
| Doing laundry | 6 (6.8) |
| Using transportation | 47 (53.4) |
| Handling finances | 15 (17.0) |

IADL: Instrumental activity of daily living

The IADL non-disabled group was significantly younger (*p*=0.001) and had a greater number of men (*p*=0.009) than the IADL disabled group. The IADL non-disabled group showed significantly higher values of UGS and IKES on both sides, KOOS-pain and PSEQ-4. Conversely, the IADL non-disabled group showed significantly lower values of PCS-6 and GDS-15 (Table 3).

**Table 3.**
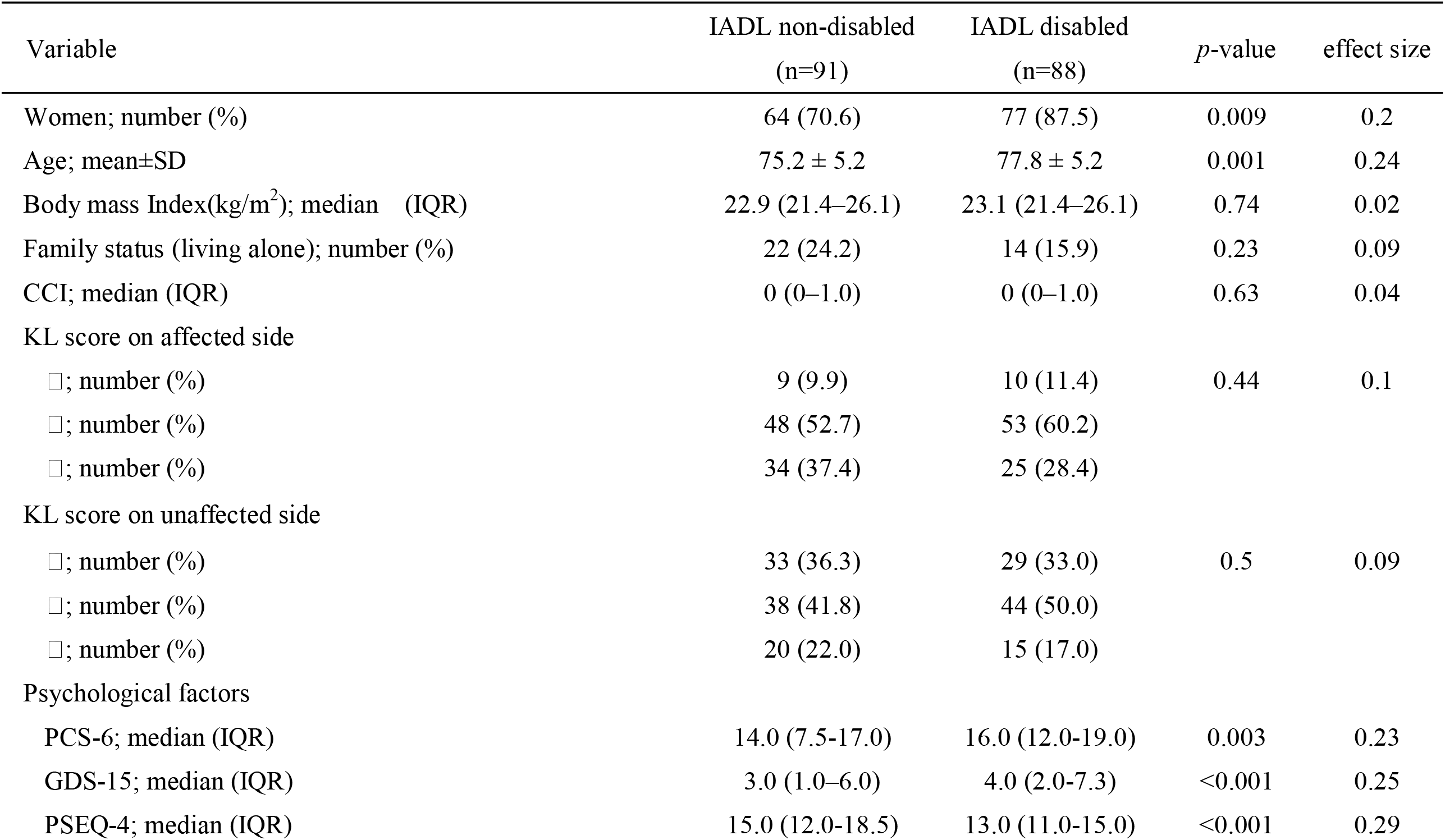
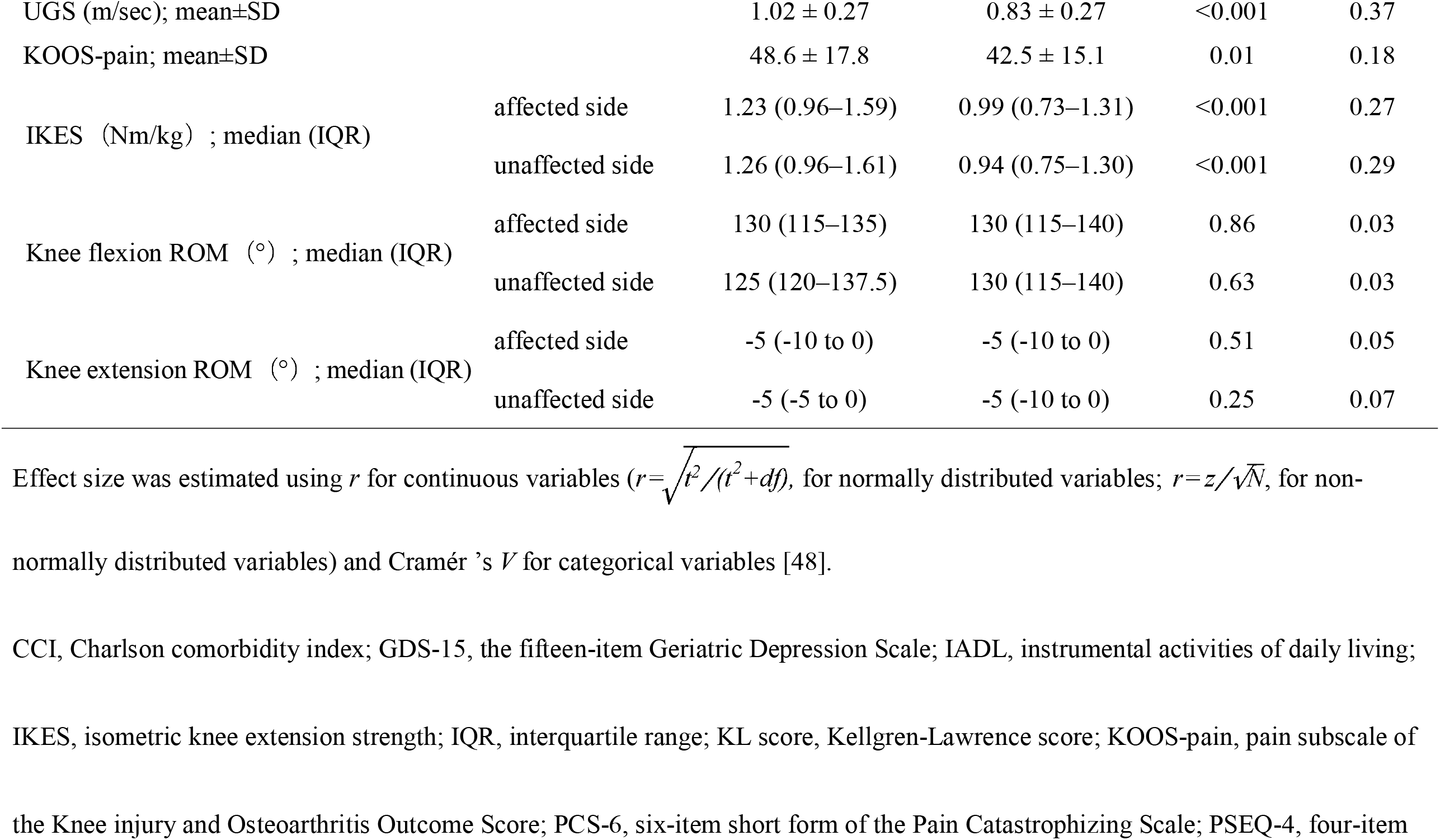
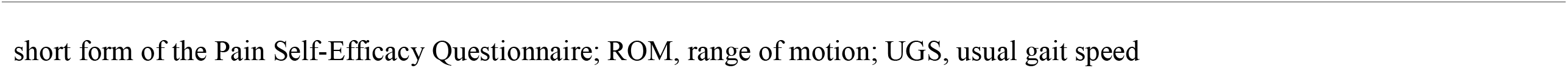
Comparison of measurements and confounders between IADL non-disabled and disabled groups.

In the binary logistic regression analysis with IADL disability status as a dependent variable, IKES on the unaffected side was adopted as a confounder. IKES on both sides showed significant differences between the two groups, but the effect size on the unaffected side was greater than that on the affected side. Therefore, only IKES on the unaffected side was included to avoid multicollinearity. In Models 1–3, PCS-6, PSEQ-4, and GDS-15 were significantly independent variables. In Model 4, PSEQ-4 (odds ratio [OR] =0.90, 95% confidence interval [CI] =0.82–0.99, *p*=0.02), UGS (OR=0.13, 95%CI=0.02–0.72, *p*=0.02), and sex (OR=0.38, 95%CI=0.15–0.96, *p*=0.04) were significantly independent variables. PCS-6 (OR=1.06, 95%CI=0.94–1.19, *p*=0.13) and GDS-15 (OR=1.06, 95%CI=0.94–1.19, *p*=0.45) were not significantly independent variables (Table 4).

**Table 4.**
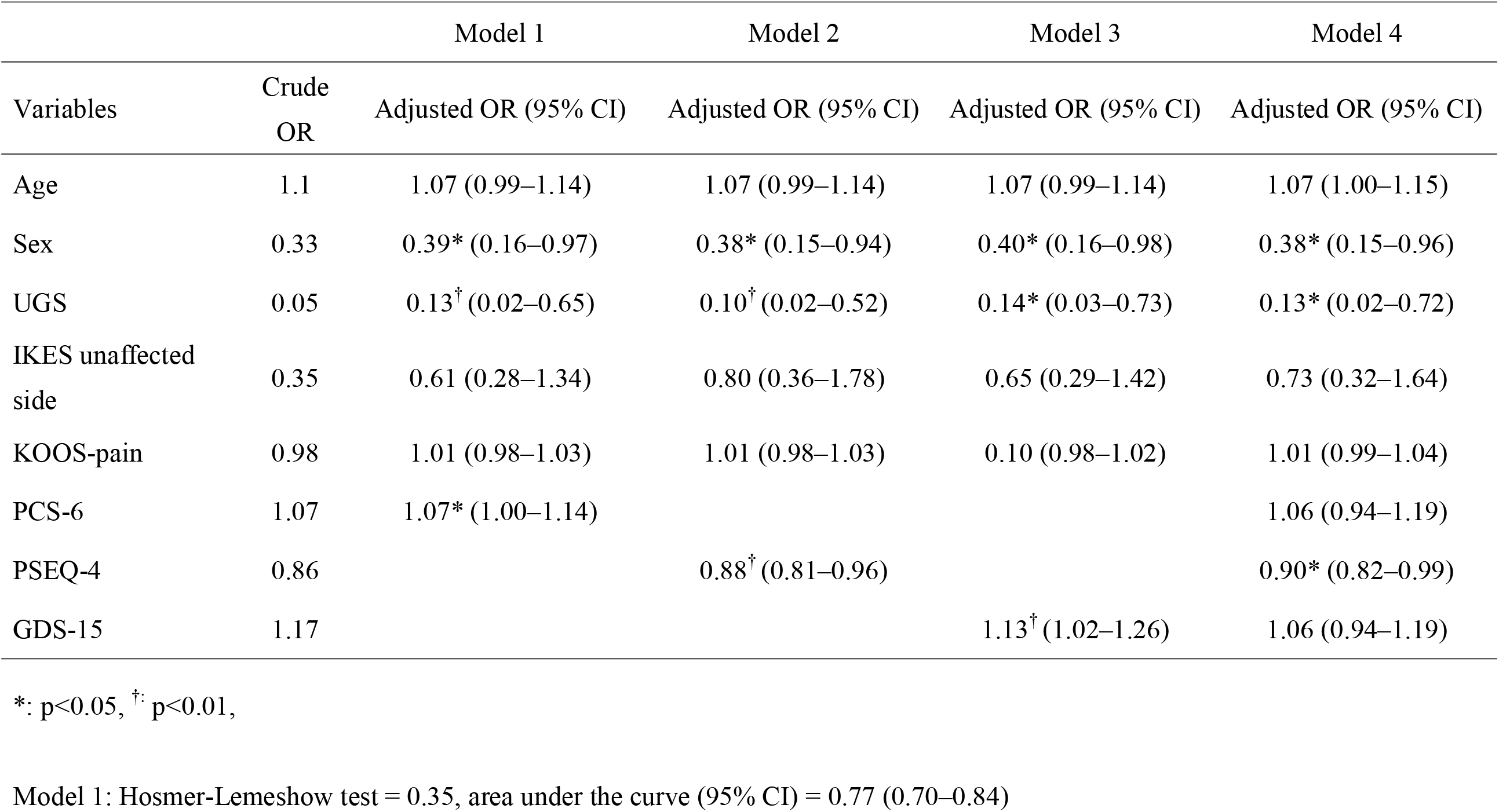
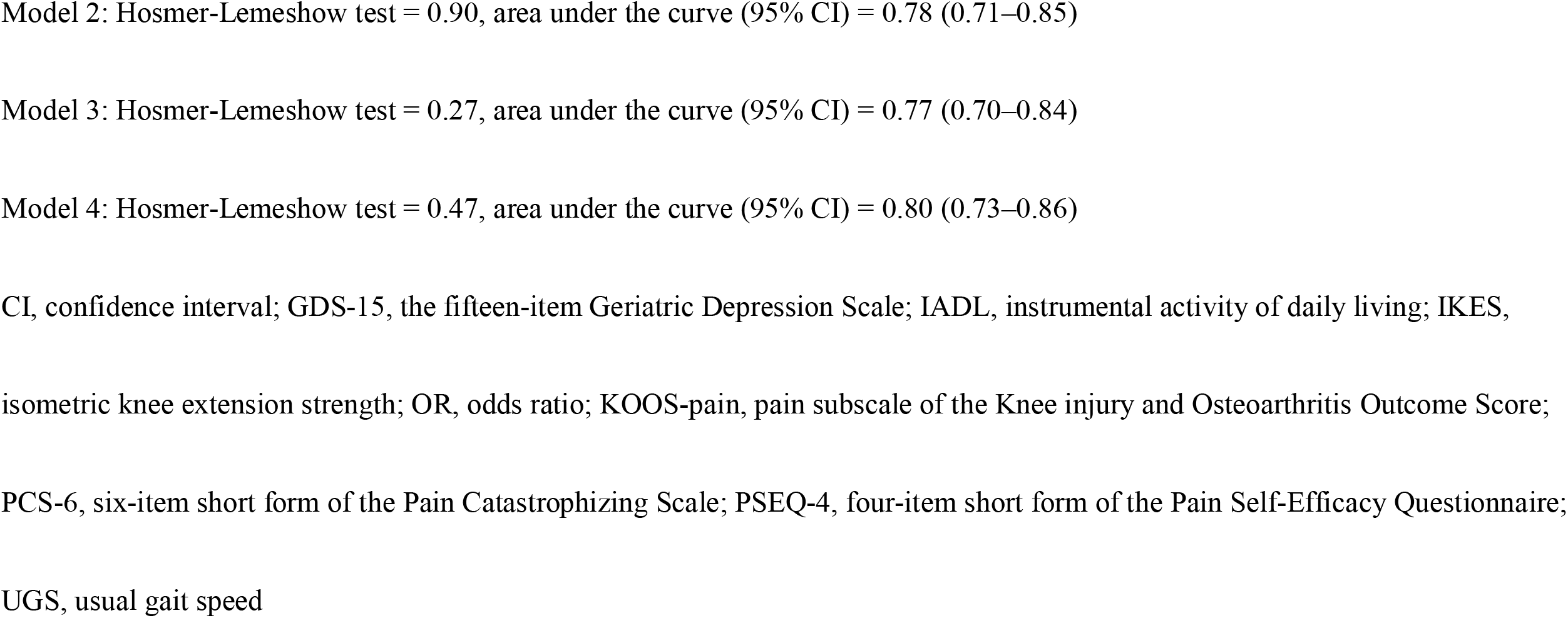
Logistic regression model with the dependent variable as IADL disabled or not.

## 4. Discussion

The present study aimed to investigate the psychological factors associated with IADL disability in older adults with moderate to severe knee OA. The values of the PCS-6, PSEQ-4, and GDS-15 tools significantly differed between the IADL non-disabled and IADL-disabled groups. However, in the logistic regression analysis, which contained all the selected psychological factors and confounders, the PSEQ-4 was identified as having significant psychological assessment capacity in association with IADL disability.

In the logistic regression model, sex and UGS were also significant independent variables. Alexandre et al. [40] have shown that sex is a factor associated with IADL disability in older adults. Our results support this finding. A reduction in gait speed is also strongly associated with IADL disability in older adults [26,33,43]. Our current findings are consistent with those of our previous study, which showed gait speed to be a discriminatory factor for IADL disability in older adults with knee OA [22]. Patients who had BADL disability (100<Barthel Index) were excluded in this study, and many participants did not have moderate to severe comorbidity assessed by CCI. However, 49.1% of all participants had IADL disability. In our study population, the main items pertaining to IADL disability were outdoor activities such as shopping (68.6%) and using transportation (50.6%), which require a degree of gait speed. Furthermore, because the effect size of UGS was the highest among all the measurements when compared between the two groups, it could be a crucial factor associated with IADL disability in older adults with moderate to severe knee OA.

Regarding psychological factors and disability in patients with chronic pain, the fear-avoidance model proposes that pain catastrophizing and depressive symptoms are some of the leading causes of disability [44]. Furthermore, self-efficacy is a mediator in the relationship between pain intensity and disability [45]. Our results implied that pain catastrophizing and depressive symptoms could be factors associated with IADL disability. However, based on the results of our multivariate analysis, self-efficacy as assessed by the PSEQ-4 was the most important factor associated with IADL disability in older adults with moderate to severe knee OA.

With respect to the relationship between pain self-efficacy and disability in patients with knee OA, Sinikallio et al. [12] suggested that self-efficacy assessed using the Pain Self-Efficacy Questionnaire was the only psychological factor associated with disability in multiple regression analysis that included demographics as confounders. Some cross-sectional studies have shown that self-efficacy, which partially includes pain elements, is a factor related to disability in patients with knee OA [14, 15]. A cohort study of older adults with knee pain reported that self-efficacy is a predictor of self-reported disability [13]. A new finding of our study is that self-efficacy assessed by PSEQ-4 was also a factor related to disability even if the disability in daily life was limited to IADL. Regarding IADL disability and self-efficacy, using a logistic regression analysis that included demographics and pain intensity as confounders, a previous cross-sectional study of older adults with chronic joint pain showed that self-efficacy is associated with IADL disability [26], which is consistent with our results. Self-efficacy describes the confidence that a person has in their ability to achieve a desired outcome [45]. In patients with knee OA, those with high self-efficacy for controlling arthritis pain have higher pain thresholds than those with low self-efficacy [46]. Therefore, older adults with knee OA who have high self-efficacy might be considered less likely to voluntarily restrict activities included in IADL, even if they have knee pain. Consequently, we considered self-efficacy, assessed using the PSEQ-4, associated with IADL disability even after adjusting for other confounders.

This study has several limitations. First, since this was a cross-sectional study, causal relationships between psychological factors and IADL disability incidence could not be determined. Second, this study included patients with knee OA scheduled for TKA or UKA, who were in a different treatment situation compared with other older adults with knee OA. Therefore, it might be difficult to generalize the results to older adults with early-stage knee OA or in a low pain status. In particular, awaiting surgery could affect pain intensity and psychological factors related to pain. Nonetheless, the average KOOS-pain score in this study population was not inferior to those in other studies [47]. The median PCS-6 and PSEQ-4 scores were also not inferior to those in previous studies in patients with chronic pain [30, 31]. Therefore, effects of awaiting TKA or UKA to pain intensity and psychological status were small. Third, because IADL disability was assessed using categorical variables as the dependent variable in our logistic regression analyses, we could not identify the effect of each factor on IADL disability. However, when comparing the two groups, the effect sizes of the variables that were significantly related to IADL disability in logistic regression models were approximately the same. This study did not aim to predict IADL disability using assessments; however, it was conducted with the conviction that greater knowledge regarding psychological factors in older adults with knee OA is likely to aid in understanding the risk of IADL disability or deterioration.

## 5. Conclusion

Regarding psychological factors, assessing self-efficacy using the PSEQ-4 was associated with IADL disability even after adjusting for confounders. Sex and gait speed were also associated with IADL disability in older adults with moderate to severe knee OA. Our study demonstrated the importance of self-efficacy, assessed using the PSEQ-4, as the factor most associated with the presence of IADL disability in older adults with moderate to severe knee OA. Future studies are necessary to elucidate the mechanism through which self-efficacy is related to IADL disability in older adults with moderate to severe knee OA. Furthermore, longitudinal studies are needed to identify predictors of the development of IADL disability in older adults with moderate to severe knee OA.

## Data Availability

The data that support the findings of this study are
available from the corresponding author, Nanjo, K., upon reasonable request.

## Acknowledgements

The authors thank D. Kurihara, K. Imahira, and K. Suda for their cooperation with data collection.

